# Comparison of Vaccination and Booster Rates and Their Impact on Excess Mortality During the COVID-19 Pandemic in European Countries

**DOI:** 10.1101/2023.03.21.23287548

**Authors:** Olga Matveeva, Svetlana A. Shabalina

## Abstract

**Aim:** To evaluate the effect of vaccination/booster administration dynamics on the reduction of excess mortality during COVID-19 infection waves in European countries.

**Methods:** We selected twenty-nine countries from the OurWorldInData project database according to their population size of more than one million and the availability of information on dominant SARS-CoV-2 variants during COVID-19 infection waves. After selection, we categorized countries according to their “faster” or “slower” vaccination rates. The first category included countries that reached 60% of vaccinated residents by October 2021 and 70% by January 2022. The second or “slower” category included all other countries. In the first or “faster” category, two groups, “boosters faster’’ and “boosters slower” were created. Pearson correlation analysis, linear regression, and chi-square test for categorical data were used to identify the association between vaccination rate and excess mortality. We chose time intervals corresponding to the dominance of viral variants: Wuhan, Alpha, Delta, and Omicron BA.1/2.

**Results:** The “faster” countries, as opposed to the “slower” ones, did better in protecting their residents from mortality during all periods of the SARS-CoV-2 pandemic and even before vaccination. Perhaps higher GDP per capita contributed to their better performance throughout the pandemic. During mass vaccination, when the Delta variant prevailed, the contrast in mortality rates between the “faster” and “slower” categories was strongest. The average excess mortality in the “slower” countries was nearly 5 times higher than in the “faster” countries, and the odds ratio (OR) was 4.9 (95% CI 4.4 to 5.4). Slower booster rates were associated with significantly higher mortality during periods dominated by Omicron BA.1 and BA.2, with an OR of 2.6 (CI 95%. 2.1 to 3.3). Among the European countries we analyzed, Denmark, Norway, and Ireland did best, with a pandemic mortality rate of 0.1% of the population or less. By comparison, Bulgaria, Serbia, and Russia had a much higher mortality rate of up to 1% of the population. Thus, slow vaccination and booster administration was a major factor contributing to an order of magnitude higher excess mortality in “slower” European countries compared to more rapidly immunized countries.

**Graphical abstract:** 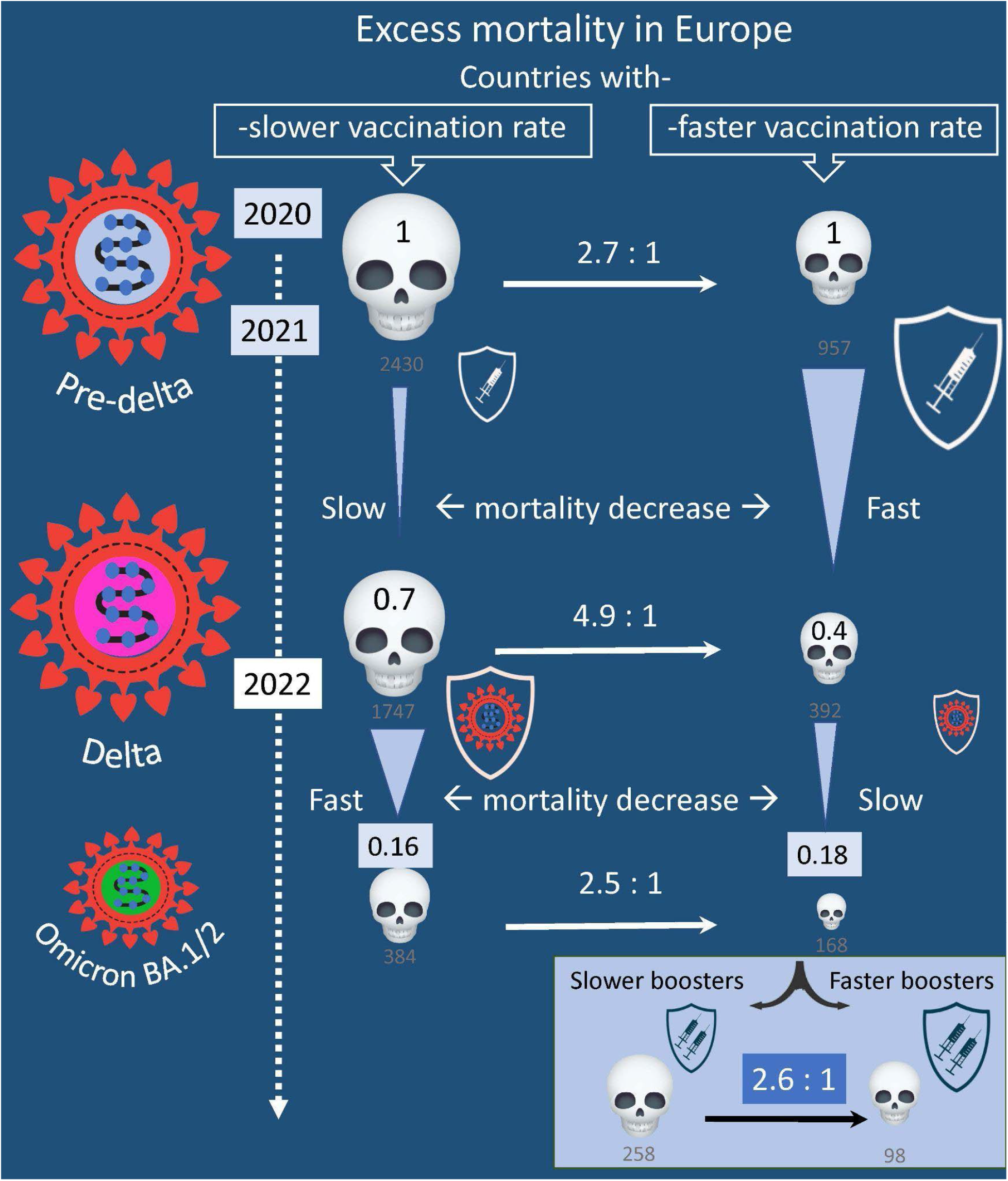

## Introduction

To deal with infectious waves caused by different variants of SARS-CoV-2, governments around the world have had to make many difficult decisions, including unpopular business closures, quarantines, and so on. An important tool in the fight against the pandemic was vaccination, which was also far from being always popular among the people of different countries and even among governments. Almost three years after the beginning of the pandemic, it is time to analyze how effective vaccination and booster administration have been during different waves of infections in various countries. One way to address this question is to compare excess mortality in different countries in which some infectious waves were synchronized, but vaccination rates varied widely. There are various vaccines that have been used to immunize populations in Europe to prevent hospitalizations and deaths from COVID-19. As shown in Supplementary Figure 1, approximately 70% of the doses administered in the EU in 2021 and 2022 are RNA vaccines produced by Pfizer or Moderna; the other 15% are non-replicating adenovirus vector vaccines produced by AstraZeneca and Johnson & Johnson. The origin of the remaining doses of vaccines administered in the EU and in other European countries according to the European vaccine tracker is unknown (1). Russia and China partially provided these vaccine portions for the EU and other European countries. For example, Russia supplied Hungary and Slovakia and predominantly vaccinated its own citizens with the adenovirus two-dose vector vaccine Sputnik V, which was not approved for use by WHO or EMA due to a lack of proper documentation (2, 3). China supplied several European countries with inactivated viral vaccines, which were approved by WHO (4). There are no studies comparing the efficacy of these vaccines back-to-back. However, many published studies have compared other vaccines used in Europe (5).

A meta-analysis of many studies shows similar effectiveness of the mRNA-based vaccines among themselves (6) and with the two-dose adenovirus-based vaccine produced by AstraZeneca (5, 7). Some studies show that the Sputnik V vaccine was just as effective (8). However, there is evidence that mRNA-based vaccines are more effective than the single-dose adenovirus vector-based vaccine produced by Johnson & Johnson (9). In addition, Lau ret al. (10) showed that virus-inactivated vaccines made in China had a shorter protection time compared with mRNA-based vaccines. It is also worth noting that several studies observed a significant reduction in the efficacy of all vaccines regarding protection against hospitalization or death caused by the Omicron virus variant compared with earlier virus variants (11, 12, 13).

In our analysis, we considered vaccination without analyzing which vaccine was predominantly used to vaccinate the country’s population. We believe that this is not a serious flaw in our study, since the vaccines used in Europe had similar efficacy. We have chosen to estimate excess all-cause mortality as a measure of the negative impact of the pandemic. This estimate can be made for countries that regularly publish all-cause mortality data. There are several models for estimating excess mortality that have been suggested for use during the COVID-19 pandemic (14, 15, 16, 17). We worked with the first of these (14) for several reasons (see below), including because its estimates are available in a very user-friendly format from the OWID database, which was the source of most of the data we analyzed. At present, however, all important findings can be substantiated by employing different models.

By comparing excess mortality estimates and other characteristics of different countries, it is possible to determine which countries did better in reducing excess mortality during each infectious wave and to try to understand why. Our study is not the first one to addresses such questions. It is consistent with others that have analyzed international heterogeneous mortality socio-economic, regulatory, and biological consequences of the COVID-19 pandemic (15, 16, 17, 18, 19). In our work, however, we focused on European countries and tried to determine exactly how much vaccination contributed to reduction of mortality.

During the coronavirus pandemic, countries around the world consistently adopted various measures to reduce mortality from COVID-19. In our work, we attempted to separate the effect that these measures had before and during mass vaccination and the effect of vaccination itself on reducing excess mortality.

Both political and biological factors influence the magnitude of all-cause mortality during pandemic infection waves (15, 16, 18). All these factors can be divided into those that change slightly and those that change or may change more radically in population during a pandemic. The first category of these factors includes the age structure of the population, GDP per capita, health care structure, and so on. The second category includes factors such as 1) the rate (ratio) of lethality in SARS-CoV-2 infections, 2) COVID-19 prevention strategies that do not include vaccination, 3) vaccination rates, 4) vaccine type, 5) population accumulation of immunity from natural COVID-19 infections, 6) the length of immune protection a person receives from a vaccine or natural infection, and 7) immune escape from the virus. Each of these factors contributes to the excess number of deaths in each infectious wave. Estimating the weight of each factor in each infectious wave is not a straightforward task. It is even more difficult to assess the causal relationship between each factor and its effect on excess mortality.

Several studies have found a negative correlation between vaccination rates and excess mortality associated with COVID-19 (20, 21, 22, 23, 24). In theory, the mere fact that such a correlation exists cannot be proof of a causal relationship. Both high vaccination rates and low pandemic-associated mortality occur in higher-income communities or countries (25, 26, 27). This inverse relationship between income and mortality was demonstrated for the 1918-1920 influenza pandemic in Europe when individual countries were analyzed (26). A similar observation was made for the pandemic COVID-19 (2020-2021). The analysis was done at the level of individual counties and zip codes (25), and globally at the individual county level (27, 28) in the US.

High GDP per capita and high vaccination rates are factors in reducing excess pandemic mortality. This is not surprising, since countries with higher income levels have more resources to deal with pandemics. Thus, they likely had more effective protective measures before mass vaccination and had higher rates of vaccination as well. There is also a collinearity between GDP and vaccination rates; a strong correlation has been demonstrated in previously published studies as well (29). Therefore, it is important to analyze how each of the factors, namely slow rate of vaccination or low GDP per capita in the population, influenced the excess mortality rate.

In our study, we tried to assess exactly how national vaccination rates were related to the mortality peaks in Europe during the Delta wave and during the first Omicron wave. European countries are leveling off in terms of excess mortality associated with waves of infections in the second half of 2022. An increase in population immunity due to natural infections probably plays a major role in this process, especially in slower vaccinated countries. Along with this process, viral immune can escape, and antigenic drift of the virus occurs relatively quickly. Even the first Omicron variants that emerged, for example, such as B.1.1.529, were characterized by an unusually large number of mutations in their spike proteins, compared with the original strain of SARS-CoV-2(30). As additional infections and booster immunizations occur, the level of protection of the population against the virus may increase, while at the same time it may decrease due to viral immune escape.

## Materials and methods

### Selection of countries

We chose European countries for our work because waves of infection caused by different variants of SARS-CoV-2 were better synchronized in Europe compared to many other regions in the world. For our analysis, we selected countries that are located entirely in Europe, except for Russia and those with a population of more than one million people. Another selection criterion was regular information updates about the coronavirus variants that were circulating in the country at any given time interval. As a result, we selected 29 countries and assigned each country a number, which we then used to refer to the country in the graphical representation of the data analysis. The countries and their assigned numbers are listed below in alphabetical order of their names: Austria 1, Belgium 2, Bulgaria 3, Croatia 4, Czechia 5, Denmark 6, Estonia 7, Finland 8, France 9, Germany 10, Greece 11, Hungary 12, Ireland 13, Italy 14, Latvia 15, Lithuania 16, Netherlands 17, Norway 18, Poland 19, Portugal 20, Romania 21, Russia 22, Serbia 23, Slovakia 24, Slovenia 25, Spain 26, Sweden 27, Switzerland 28, United Kingdom 29. Almost all these countries are members of the European Union. The exceptions are Norway, Sweden, UK, Russia, and Serbia. The names of the countries that fall into each category are shown in Supplementary Figure 1.

### Datasets

Actual GDP per capita data were retrieved from the World Bank dataset (31). Vaccination and mortality values were taken from the Our World in Data (OWID) project dataset (32, 33). The time intervals when different variants of the coronavirus dominated were calculated from the information provided in the OWID database.

### Characteristics of the countries

Collected country characteristics for each analyzed time interval were recorded in an Excel file available as a supplemented material.

### Vaccination

For each selected European country, we retrieved from the OWID database the percentage of fully vaccinated people for several time points. For the countries included in our analysis, full vaccination usually means receiving a primary series: two doses for vaccines with a two-dose course and one dose for vaccines with a one-dose course. We retrieved this percentage from the database for three dates: July 2, 2021, October 2, 2021, and January 2, 2022. We also collected information on how many people in each country got booster doses of the vaccine by January 2, 2022. A booster dose refers to a third or fourth injection of the vaccine. The OWID database does not tell you whether the booster dose was given once or twice.

### Mortality

We extracted estimates of confirmed COVID-19 deaths from the OWID project database, as well as estimates of excess deaths over various pandemic time intervals. The latter estimates were obtained using the Karlinsky-Kobak model. The model algorithm was created first by fitting a regression model for each country using historical mortality data for 2015-2019, second by using the resulting model to predict the number of deaths that can be expected in 2020-2022, and third by subtracting expected deaths from those reported for each country (14).

Mortality estimates were obtained for the three time periods labeled I, II, and III in the graphical presentations of our analysis. The first of these periods (I) is the time interval before the start of mass vaccination (February 2020-June 2021). The second (II) period (July 2021-January 2022) and the third (III) period (February 2022-May 2022) correspond to the time when vaccination was in full swing.

Two virus variants, namely Wuhan and Alpha, dominated the first period in sequential order, Delta dominated the second time interval, and Omicron BA.1 and BA.2 dominated the third. The precise data collection time points of the Delta/Omicron dominance switch in January or February were defined individually for each country. For some countries, this switch occurred in January and for some in February 2022. Technically, using OWID available information, we determined the date when the Delta/Omicron balance in a country approached 50% and estimated excess mortality or mortality from COVID-19 in that country after three weeks from this date. We assumed that one week after the 50% point of presence in the country, Omicron dominates. Thus, we chose the time point at which Omicron dominates, and we assumed that the mortality is mainly due to the infection associated with this strain two weeks after the corresponding date. In other words, three weeks after the date when Delta and Omicron accounted for half of the infections, we assumed that deaths occurred mainly from Omicron infection.

### Categorization of countries

We categorized countries according to their vaccination rate as “faster”, or “slower”. The “faster” category included countries that had reached 60% of vaccinated residents by Oct. 2, 2021, and 70% by Jan.2, 2022. The “slower” category included remaining countries that had not reached these levels of vaccination during the relevant time intervals. In the “faster” category, a subcategory with a vaccination rate of 35% achieved as of January 2, 2022, or more, was created and named “boosters fast”. We show the names of the countries that fall into each category in Supplementary Figure 1.

### Analysis

Pearson correlation analysis, linear regression fitting, and Odds Ratio calculations were performed using Excel calculation tools. We verified our results of categorical Chi-Square test calculation by using online tools (34, 35).

## Results

### Comparative analysis of GDP, mortality, and vaccination rates in European countries

In this study, we were most interested in the relationship between COVID-19 mortality and vaccination rates. However, when examining this aspect, we need to consider that countries differ not only in vaccination rates but also in other characteristics that may be related to population survival in a pandemic. For example, one of the most important characteristics of a country is the actual GDP per capita. This parameter can correlate strongly with the level of health care and the effectiveness of pandemic mitigation strategies, which are directly related to the reduction of excess deaths.

If a country has a small GDP, it is more difficult to implement anti-epidemic measures and keep the economy afloat. European countries are extremely diverse by this criterion: their GDP per capita varies by orders of magnitude: from $9,200 in Serbia to more than $90,000 in Switzerland and Ireland (Figure 1 A).

**Figure 1.**
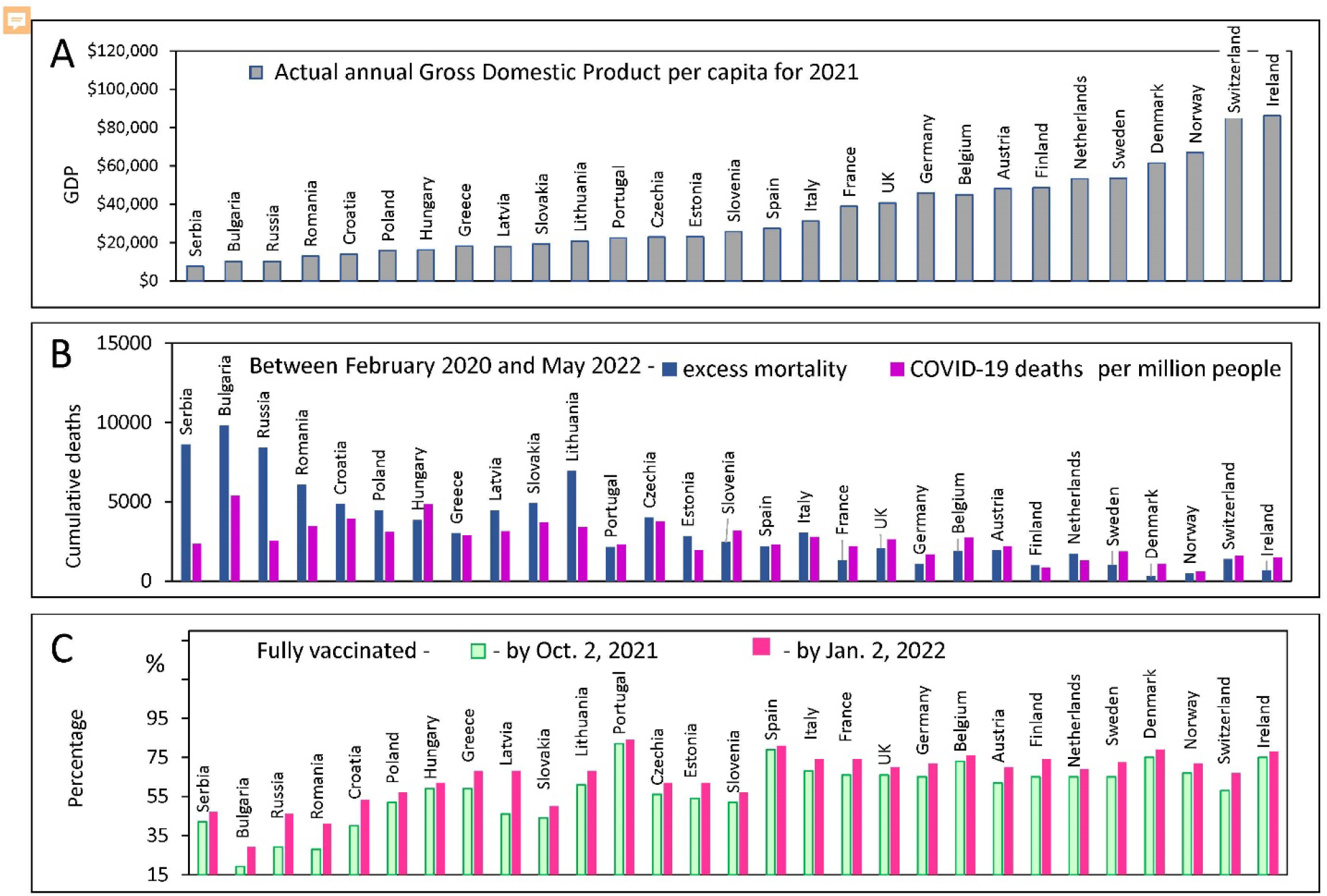
Country characteristics: GDP, Mortality and Vaccination Rates. Countries are ranked in order of increasing GDP per capita in 2021 in all panels. Standard deviations and error bars are not shown. A. Actual annual GDP values from the World bank database for 29 European countries. B. Excess mortality estimates, and COVID-19 confirmed deaths from the OWID database C. The percentage of people who received two doses of vaccine (primary series) at certain dates.

We investigated how the diversity of annual income relates to the diversity of other countries’ characteristics of interest. We were particularly interested in COVID-19 mortality rates, the excess mortality rate during the pandemic, and the vaccination rate achieved in a country at different intervals. These characteristics are shown in Figure 1.

In the same order as national per capita GDP growth, Figure 1B demonstrates the number of confirmed COVID-19 deaths and estimated excess mortality during the pandemic. In addition, in the same order of countries, Figure 1C also shows national vaccination rates.

Visualizing country characteristics reveals several trends. Countries with relatively low GDP per capita (less than $15K) tend to have COVID-19 mortality rates that are significantly lower than the excess all-cause mortality estimates (Figure 1B). In addition, these same countries with lower GDPs have relatively low vaccination rates (Figure 1C). The discrepancy between COVID-19 deaths and excess deaths may indicate a problem of deficiencies in disease diagnosis. A similar discrepancy was observed in several other studies published previously (17, 36).

Given the problem of possible under-reporting of COVID-19 deaths in some countries, we decided to bypass the ambiguity of confirming such deaths and analyze the data solely with excess mortality estimates.

### Negative correlation between GDPs per capita and excess mortality estimates

Trends that hint at the relationship between per capita GDP and mortality during the pandemic, as well as the level of vaccination achieved by the country, were explored in more detail in the next step of our analysis. This analysis showed a significant negative correlation between excess mortality and a country’s actual GDP per capita. The correlation is much more pronounced for poorer countries - those with a GDP per capita of less than 40K. It is much weaker for richer countries (Figure 2A). For the category of countries with GDP under 40K, the correlation is highly significant (R=-0.79, p<<0.01), and for the others, it does not reach the 95% level of significance (R=-0.53, p=0.08).

**Figure 2.**
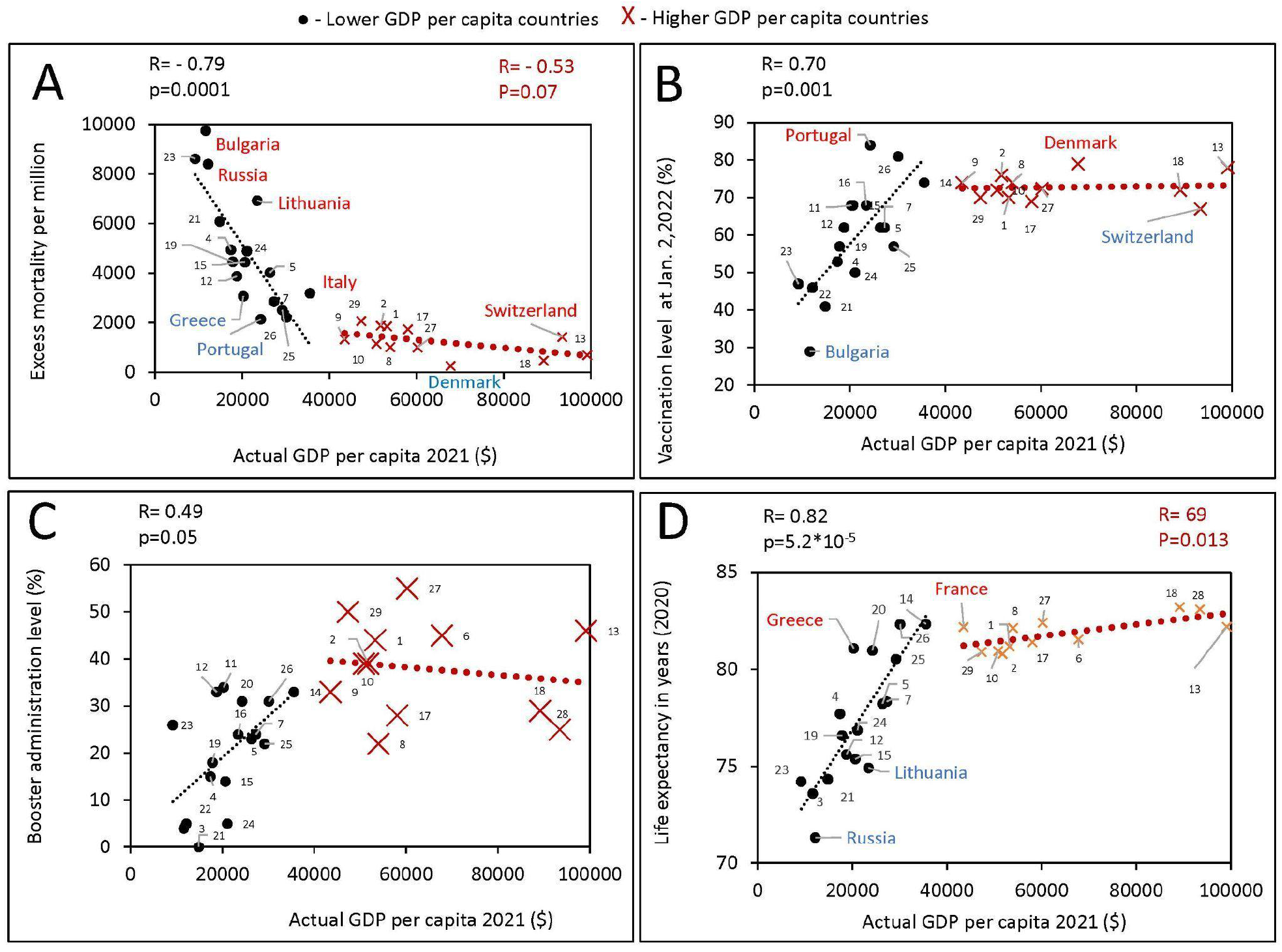
Relationship between GDPs per capita and various characteristics of European countries obtained during different periods of the pandemic. All estimates of excess mortality in the scatter plots are per million population. Higher GDP per capita countries were defined in this study as those with a GDPs equal to or greater than the threshold of $40,000, lower GDPs per capita countries defined as those with a GDPs per capita below the threshold. Countries were numbered according to the alphabetical order of their names as follows: Austria 1, Belgium 2, Bulgaria 3, Croatia 4, Czechia 5, Denmark 6, Estonia 7, Finland 8, France 9, Germany 10, Greece 11, Hungary 12, Ireland 13, Italy 14, Latvia 15, Lithuania 16, Netherlands 17, Norway 18, Poland 19, Portugal 20, Romania 21, Russia 22, Serbia 23, Slovakia 24, Slovenia 25, Spain 26, Sweden 27, Switzerland 28, United Kingdom 29. A. Excess mortality during the COVID-19 pandemic from February 1, 2020, to May 30, 2022, in relation to actual GDP per capita as of 2021. B. Vaccination rates, as of October 2, versus actual GDP per capita. C. Boosters administration rates, as of January 2, versus actual GDP per capita. D. Life expectancy versus actual GDP per capita.

It is worth noting that among countries with incomes under 40K, Lithuania, Bulgaria, Italy, and Russia lost far more people than countries with close or the same incomes, while Greece and Portugal lost far less (Figure 2A). At the same time, among high-income countries (above 40K) Switzerland stands out negatively as losing more people, while Denmark stands out positively because it lost less. This conclusion can be drawn because the first group of the countries is above the corresponding trend line, while the second group is below it.

### Positive correlation of GDPs per capita with rates of vaccination and booster administration as well as life expectancy

We found a positive correlation between GDPs per capita and vaccination rates in countries with a GDP less than 40K, as shown in the scatterplot in Figure 2B. No such correlation was observed for countries with higher GDPs (Figure 2B). It is worth noting that Portugal and Denmark have higher vaccination rates, while Switzerland has lower vaccination rates than the trend suggests. Interestingly, during the COVID-19 pandemic, Bulgaria was the country that lost the most lives per million population, while Denmark was the country that lost the least lives among the European countries we analyzed. Bulgaria had the lowest vaccination rate among countries with the same income level, and Denmark had the highest.

As with the patterns identified earlier, we observe a pronounced correlation between the level of boosters’ administration and country income for those countries with GDP per capita below 40K (Figure 2C). However, it is worth noting that even in these lower-income countries, the correlation between GDP and the percentage of the country’s population that received a booster dose of vaccination is relatively weak *R*=0.49, *p*=0.05; in richer countries, it disappears altogether.

There is a very pronounced positive correlation between GDP and life expectancy for countries with a GDP of less than 40K (*R*=0.82, *p*<<0.01). In contrast, a weaker correlation exists for countries with higher income levels (*R*=0.69, *p*=0.013). Greece and France stand out positively, with a higher life expectancy than in countries with the same income level. While Russia and Lithuania stand out negatively, people in these countries live less than in countries with the same income level.

### Time intervals, estimates of excess mortality and dominant virus variant for COVID-19 infection waves

Coronavirus infection has spread in Europe in several waves caused by different variants of SARS-CoV-2. As reported smoothened weekly COVID-19 deaths, these waves are shown in Figure 3A. The first wave, presented in Figure 3A, shows deaths caused by the ancestral Wuhan variant of the virus. The second large wave with several peaks in the graph of Figure 1A represents deaths caused mainly by SARS-CoV-2 Alpha variant, which was dominant in Europe before the Delta variant emerged. The next large wave with two peaks shows deaths during the time interval when Delta and Omicron BA.1/2 variants dominated. Finally, the last visible comparatively small waves represent COVID-19 deaths that occurred during the time intervals when Omicron BA.5, BQ.1, and XBB 1.5 dominated.

**Figure 3.**
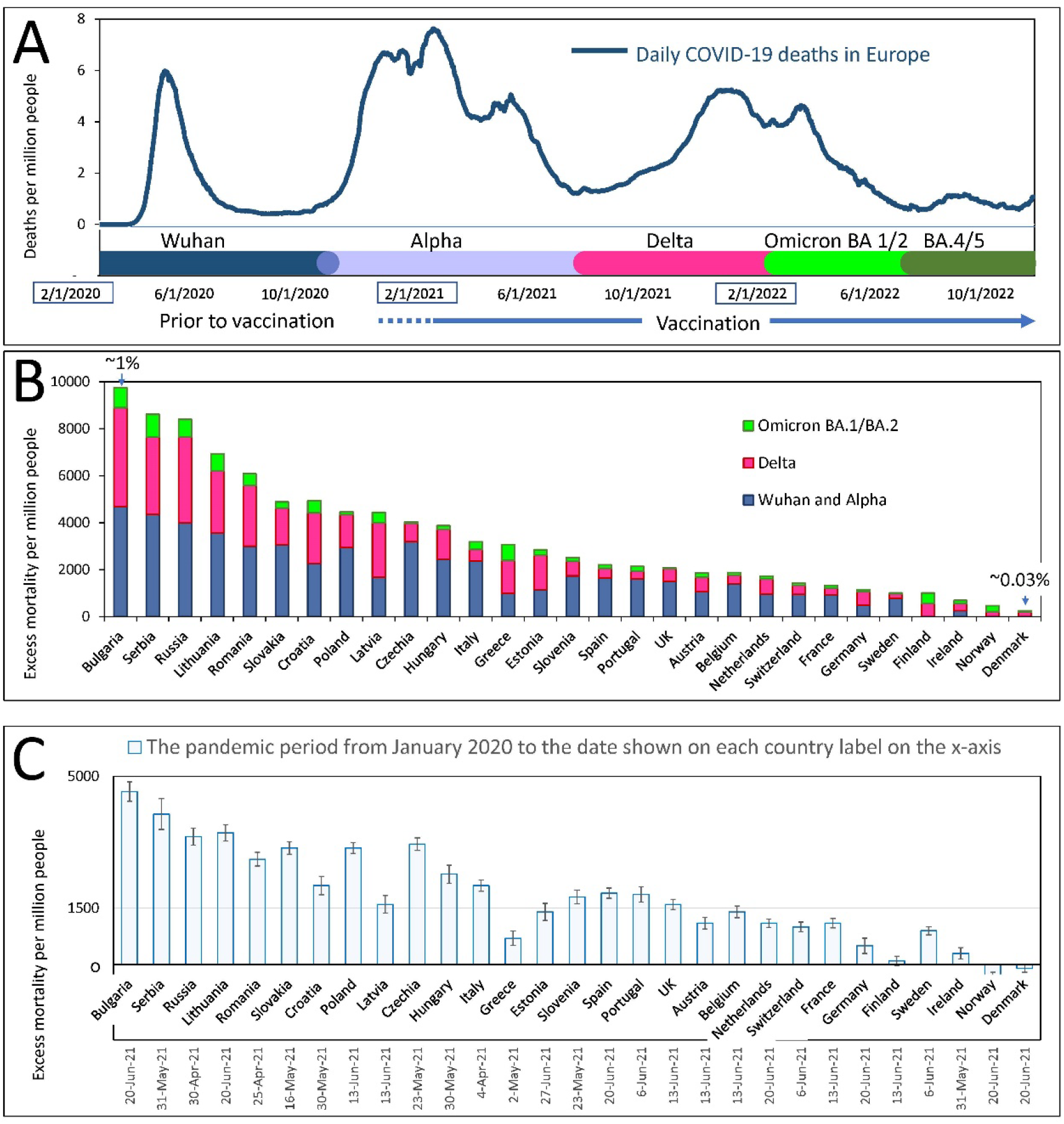
Mortality during COVID-19 pandemics in Europe. A. Visualization of infection waves as weekly averaged confirmed COVID-19 deaths in Europe during the pandemic through October 15, 2022. Different time intervals are highlighted in distinct colors. Information on excess mortality and dominant virus variants in each time interval was taken from the OWID database. The information is shown in more detail in the Supplementary Figure 1. B. Excess mortality estimates for each European country for the COVID-19 pandemic period from February 2020 to May 2022. Estimates are from the OWID database and are derived from the Karlinsky-Kobak model. Each column represents three different time intervals, which are highlighted with the same colors as in “A”. C. Karlinsky - Kobak model estimates of excess mortality values over the time from the beginning of the pandemic up to the summer of 2021 with the standard deviation intervals.

In Europe, as noted above, the Delta-driven wave and the Omicron BA.1 wave were very close to each other and appeared as one big wave with two peaks (Figure 3 A). However, at the level of each country, these waves were often separated from each other. By analyzing the individual data for each country, we believe we could determine the time interval at which people mostly died from the severe course of infections caused by a particular variant of the virus.

The overall estimates of excess mortality, which range from a few hundred to a few thousand per million people in each country, show a contrast in the impact of the pandemic on European countries (Figure 3B). At the extremes of the spectrum, illustrating this contrast, are countries that have lost less than one-tenth of their residents and countries that have lost nearly one percent of their citizens’ lives. Thus, we can conclude that some countries have handled the pandemic better than others, reducing the excess deaths. Among the European countries we analyzed, Denmark, Norway, and Ireland handled the pandemic best. The worst coping countries that suffered the greatest losses of lives were Bulgaria, Serbia, and Russia.

Figure 3B shows that not only did countries differ in excess mortality throughout the entire pandemic period, but differences were observed in each time interval corresponding to each infectious wave. In the following section, we analyze the data more thoroughly to understand what factors led to these differences.

The excess mortality estimates presented in Figure 3B were calculated using the algorithms of the Karlinsky-Kobak model (14). To avoid presenting too much detail in Figure 3B, we have provided estimates of the standard deviation (SD) of the model calculation separately in Figure 3C.

Some countries showed negative excess mortality values. This effect can probably be explained by lockdowns, quarantine, which reduced the number of car accidents, and social distancing, as well as mask-wearing, which reduced the prevalence of influenza and other viral infections that can affect national mortality.

### Comparison of excess deaths that occurred early and late in the pandemic

COVID-19 vaccination began in late 2020, but by February 2021, only two percent of the population in Europe had been vaccinated. It is interesting to compare the number of COVID-19-related casualties before mass vaccination began or would have had a protective effect in European countries and after. We chose July 2, 2021, as the date to separate the two periods. By this date, most European countries had vaccinated over 8% but less than 40% of their fellow citizens. We present this information in more detail in Supplementary Figure 2.

Until July 2021, the Wuhan and Alpha virus variants prevailed in Europe, and after that month, the Delta variant, followed by Omicron. Thus, most of the excess deaths before July 2021 happened before vaccination protected people in masse. The periods we compared differ both in the level of vaccination and in the type of dominant viral variant. Wuhan and Alpha dominated for almost a year and a half and claimed a certain number of lives. Delta dominated for six months and claimed many more lives in some countries than previous variants of the virus over the same period. In other countries, Delta claimed far fewer lives. The question is - why?

To answer this question, we compared the number of deaths in each country before and after mass vaccination. This analysis allowed us to understand how well countries protected their populations in the periods separated by July 2021. The results are presented in the two scatterplots in Figure 4. The top plot in Figure 4 shows that there is a significant correlation between the rates of excess mortality that occurred during the two periods of interest (R=0.82, p<<0.001). Thus, some countries, such as Bulgaria, Serbia, and Russia, lost the most lives during both time periods of interest. Similarly, some countries, such as Finland, Norway, Denmark, and Ireland, had the fewest deaths in both time periods.

**Figure 4.**
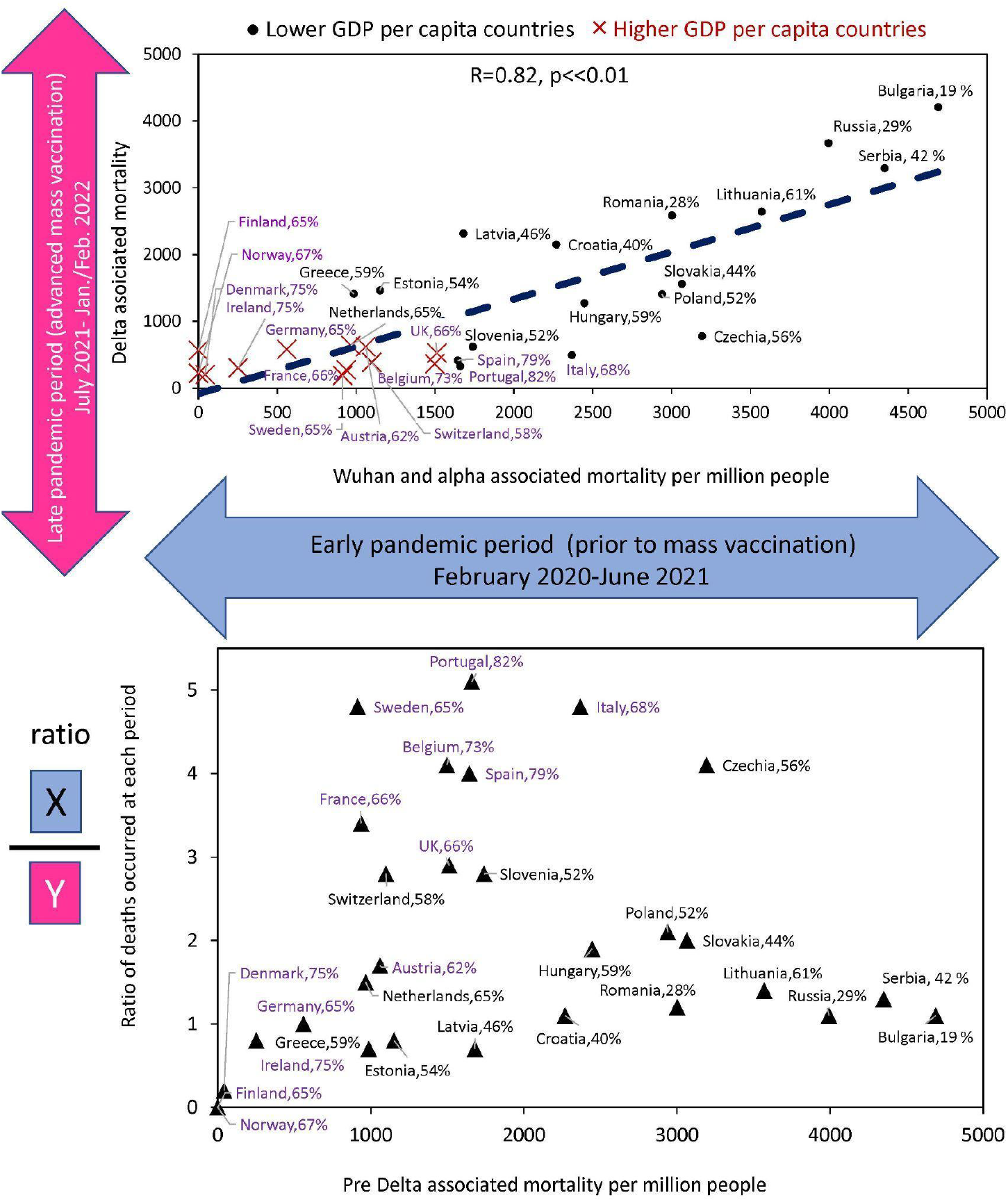
Relationship between estimates of excess deaths occurring earlier and later during the COVID-10 pandemic. The top graph shows the correlation between deaths that occurred in the country during Delta domination relative to deaths whose culprits were pre-Delta virus variants (Alpha and Wuhan). The lower graph shows the ratio of deaths in the two periods for the same countries. All names of countries that fall into the “faster” vaccination category are highlighted in lilac font. The names of other countries are in black. The percentage of the vaccinated population that was completely in the country by the time Delta arrived is shown after the name of the country.

It is worth noting that countries below the trend line in the upper plot tend to have higher vaccination rates than x-matching countries above the trend. This indicates to us that those with higher vaccination rates reduced mortality in the Delta wave more than those with lower vaccination rates.

The bottom graph in Figure 4 shows the ratio of deaths between the two periods for each country relative to deaths during the first period. It illustrates how effectively countries have improved the protection of the lives of their citizens in the second period compared to the first. The contrast is staggering! Portugal, Sweden, Italy, the Czech Republic, and Belgium reduced mortality by a factor of four or more, while Serbia. Bulgaria, Russia, and Lithuania had virtually no reduction in losses. Thus, some countries, after experiencing high mortality in the first period, radically changed their trajectory and protected their populations much better in the second period, when faced with a Delta variant of the virus. Is the change in trajectory related to rapid vaccination? The answer to this question is yes. Indeed, most of the countries with rapid vaccination rates are among those that have significantly reduced mortality rates during the Delta-dominated period. At the same time, countries with very low vaccination rates have increased their mortality rates.

### Correlation analysis of excess mortality estimates versus vaccination rates

In the next step of our work, we analyzed the estimated excess mortality values in more detail in the context of factors that might affect COVID-19-related mortality. Towards this goal, we created scatter plots showing estimates of excess mortality over time as a function of vaccination/boosters rates. Each country was represented by an excess mortality value that was plotted along with vaccination or booster administration rates, achieved at specific time points in the same countries (Figure 5).

**Figure 5.**
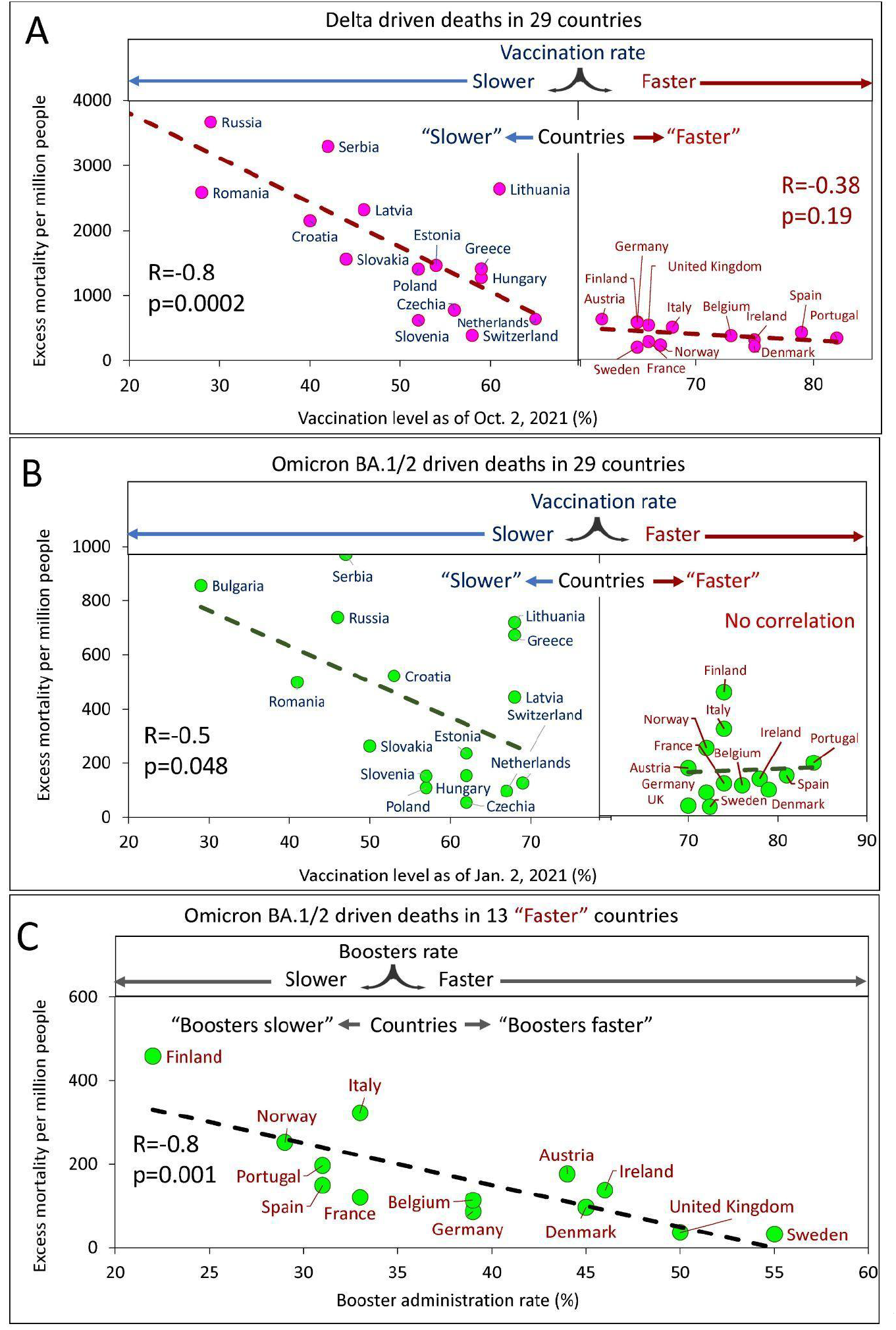
Excess mortality and vaccination rates in European countries estimated for different time intervals. A. Delta associated excess mortality by country vs. vaccination rates. B. Omicron BA.1/2 associated excess mortality by country vs. vaccination rates. C. Omicron BA.1/2 associated excess mortality by country vs. boosters’ administration rate in the “faster” vaccination rate country category.

Analysis of the data showed that a higher vaccination rate in a country corresponded to a lower excess mortality rate. By sampling the data and linear interpolation between points, we found that the best linear trend of decreasing mortality as the vaccinated population in a country increased was in countries with vaccination rates below a certain threshold. This threshold is somewhere between 60 and 70% of the vaccinated population in the country. Thus, we found that for countries with vaccination rates below the threshold; the correlation is very strong and significant, while for countries with vaccination rates above the threshold, the correlation is weak, marginal (Figure 5A) or virtually not-detectable (Figure 5B). Low excess mortality, which was not significantly different between countries, was observed in all countries with vaccination rates above the threshold. This observation was accurate for the Delta-dominated period (Figure 5A) and for the Omicron BA.1/2 dominated period as well (Figure 5B). Such an analysis of the data allows us to divide the countries into two categories: those that vaccinated faster and those that vaccinated slower. The countries with higher vaccination rates reached some threshold level and equaled them in terms of excess mortality rates.

At the same time, for the Omicron BA.1 and BA.2 period, we found that in countries in the “faster” vaccination category, while mortality was practically independent of vaccination rate, it depended on booster rate (Figure 5C).

We can summarize these results as follows. A significant negative linear relationship between excess mortality and vaccination rates was found only for “slower” countries. This was true both for the Delta dominant period (*R*=-0.8, *p*<<0.01) and for the Omicron BA.1 and BA.2 dominant period (*R*=-0.5, *p*=0.045). For countries in the “faster” category, we did not find significant relationships. All countries in the “faster” category have relatively low mortality rates. However, an inverse linear relationship between mortality and the rate of booster administration was found for the “faster” countries during the Omicron-dominated period. The greater the percentage of the population that received booster doses, the lower the mortality rate (*R*=-0.8, *p*<<0.01).

Far more people died from COVID-19 in countries that were slow to vaccinate their populations compared to countries that did it faster. We can say the same of the speed at which countries provided additional booster vaccine doses. The higher the speed, the fewer deaths.

### Age characteristics and excess mortality

We were interested in the question of how a country’s age characteristics relate to excess mortality. To conduct an analysis that would answer this question, we chose three characteristics, namely the average age of the population in the country, the percentage of the elderly population (65+), and life expectancy. It should be noted that of the three characteristics listed, only life expectancy of the population is the most related to the quality of life and health care. The average age and the percentage of the elderly population (65+) are less dependent as they are strongly influenced by the percentage of young people in the country. We examined these three characteristics and their correlation with excess mortality,

Figure 6 illustrates the results of the analysis, which correspond to infection waves caused by Delta and Omicron virus variants BA.1/BA.2. There is no significant correlation between the average age of a population and excess mortality in a country (Figure 6A). The lack of a significant correlation is also seen in panel B of Figure 6, which shows the percentage of the elderly population and excess mortality in the country. However, we can see a pronounced and significant negative correlation between life expectancy and excess mortality (Figure 6C). The longer a life expectancy in a country, the lower the pandemic excess mortality. This is true for both the Delta wave and the Omicron wave.

**Figure 6.**
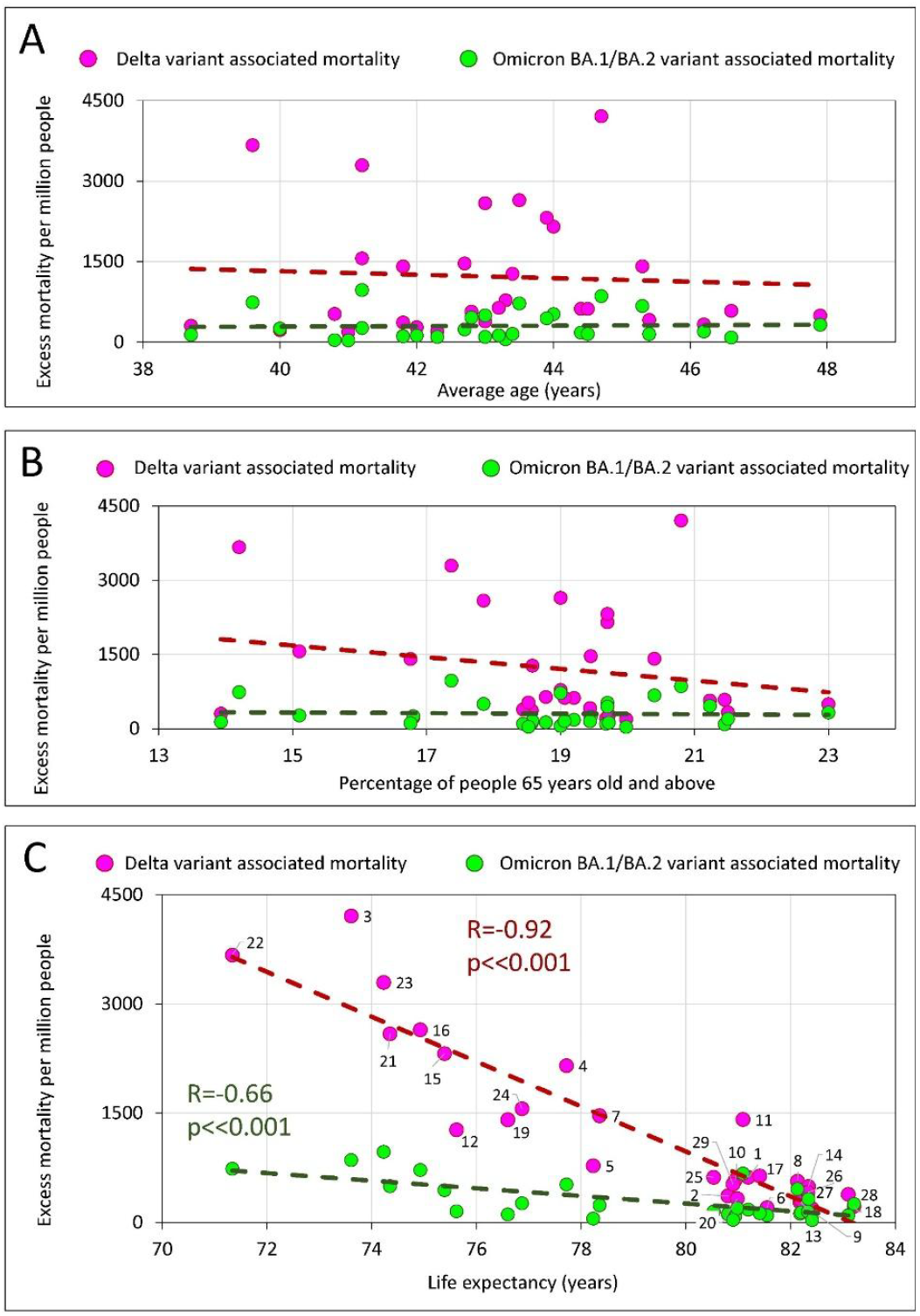
Excess mortality and the average age of the population in the country, the percentage of the elderly, and life expectancy in the country. A. Excess mortality and the average age of the population in the country. B. Excess mortality and the percentage of the elderly population (65+) in the country. C. Relationship between excess mortality and life expectancy.

We conducted the same correlation analysis with the same parameters separately for the category of countries that vaccinated their populations faster and for the category of countries that did so slower (Supplementary Figure 3). In this separate analysis, we see that there is a trend in countries that vaccinated more rapidly, showing that the higher the average national age, the higher the excess mortality. The trend is there, but it does not reach statistical significance (*R*=0.47, *p*=0.09). However, we found no correlation between the percentage of the elderly population and excess mortality in a country, either when analyzing all countries together or when analyzing the categories of countries in which the population was vaccinated with different rates (Supplementary Figure 3, middle top, and middle bottom panels).

### GDP per capita and vaccination rate as inputs for regression models

In trying to establish a causal relationship between vaccination rates and the reduction of excess mortality, it is important to understand what factors other than vaccination contribute to saving the lives of the nation’s citizens. Some of these factors might be related to GDP. This is not a straightforward task, because the level of a country’s GDP itself is correlated with the level of vaccination. We demonstrated the existence of this correlation in our study (Figure 2B) and the result is consistent with the results of the previously published research (19, 29).

To address this issue, a set of linear regression models with variable input parameters was created. In all these models, mortality was used as the dependent variable, and per capita GDP values and countries’ vaccination or boosters’ rates were treated as independent variables. The results of linear regression analysis, as the columns showing the level of significance that each model assigned to each parameter, are presented in Figure 8. More information about the results of the models is available in Supplementary file 1.

We found that to predict excess mortality, depending on the time interval when the deaths occur, either the levels of GDP or the vaccination rate or both may be significant as input parameters (Figure 7A). Model 1, which considers mainly COVID-19 mortality in the period before mass vaccination and before the dominance of the Delta variant, assigned a level of significance only to the GDP parameter. In contrast, Model 2, which considers mortality that occurred when the vaccination process was advanced and the Delta variant dominated, assigned significance levels to both input parameters: namely, GDP and vaccination. Finally, model 3, which analyzes mortality during the period of advanced vaccination, when the Omicron variant dominates, assigns significance levels only to the vaccination rate. It should be noted that the vaccination parameter appears to be much more significant in Model 2 compared to Model 3.

**Figure 7.**
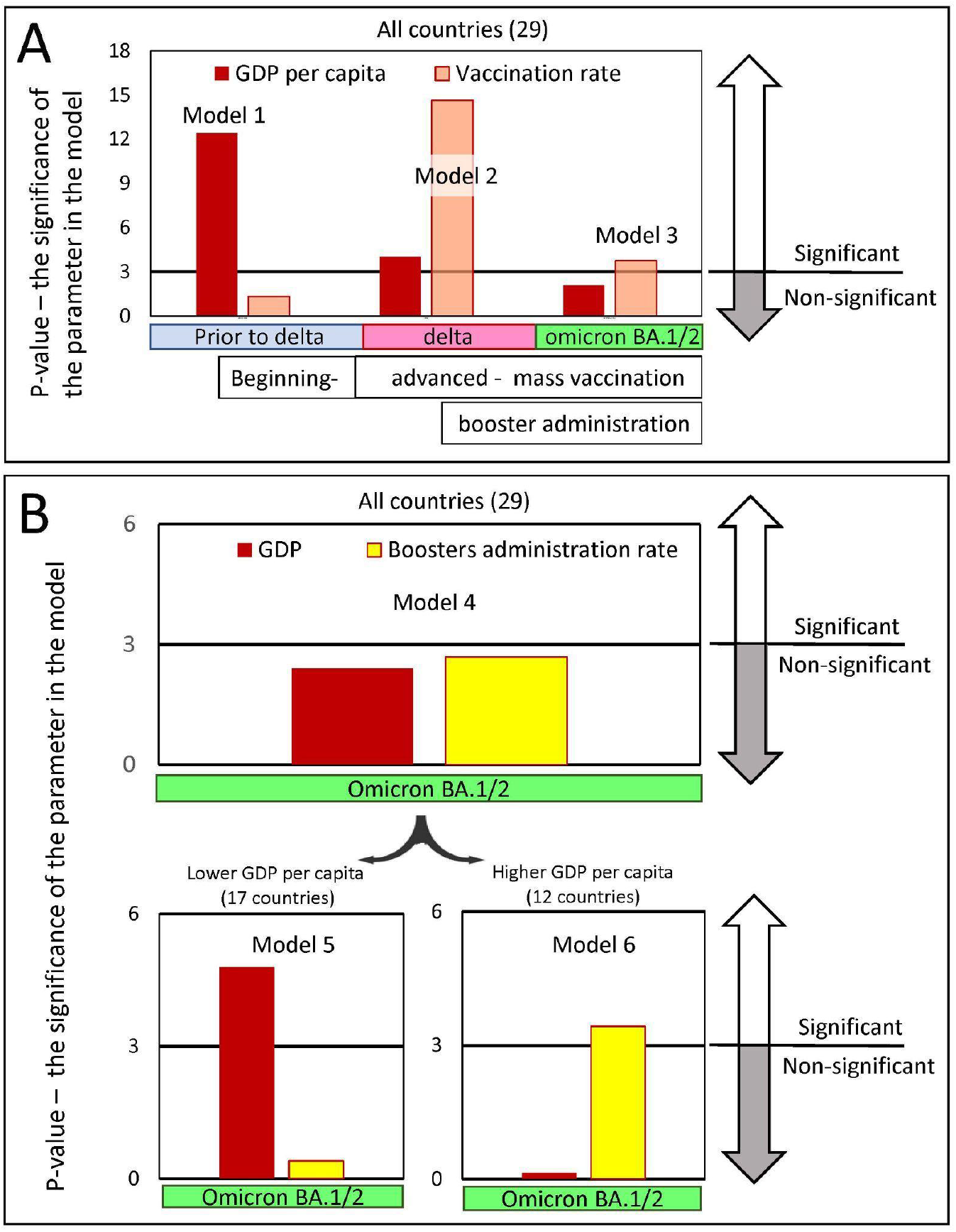
Significance levels of input parameters in linear regression models predicting pandemic excess mortality. Logistic regression models were created to predict excess mortality. Mortality was chosen as the dependent variable, while GDP per capita and the vaccine immunization rate of the country’s population were chosen as the independent variables. Each figure column represents a negative logarithmic value of the significance level of the corresponding model input parameter. The pandemic periods, for which we built models, differed in the dominance of the viral variant. Wuhan and Alpha variants dominated Europe, before July 2021, afterwards the Delta variant dominated until January/February 2022 and later until May 2022 Omicron BA.½. A. Input parameters are GDP and vaccination rates in the country, achieved in the respective time periods. Models 1-3 were created for the set of all 29 countries. B. Input parameters are GDP and the level of boosters in the country reached by a certain time. Model 4 was created for the set of all 29 countries, and models 5 and 6 were created only for countries subsets with GDP per capita below or above 40K, respectively. There are 17 countries in the first subset and 12 in the second.

Thus, summarizing the results, we can conclude that the factors affecting mortality reduction related to the country’s income played the greatest role before mass vaccination began, but the least afterward. We can also conclude that the level of vaccination played a greater role in preventing Delta-induced deaths compared to Omicron-induced deaths.

It is interesting to examine models that consider GDP and the level of boosters achieved in the country as input parameters by the time the Omicron virus variant emerged. We present the results of such studies in Figure 7B. Looking at the results of Model 4, which shows an analysis of all countries, one might get the impression that neither GDP nor booster levels play an important role in mortality in a country during Omicron dominance. This conclusion can be drawn because none of the relevant parameters are significant as inputs to Model 4. This conclusion is flawed, and this becomes clear if we separately analyze the categories of countries with lower or higher GDP per capita.

For countries with relatively low income, it is this parameter of GDP that is most significant in determining high mortality (Figure 7B, model 5), while, by contrast, for countries with higher income, GDP is not significant in the model, but the booster administration rate is highly significant (Figure 7B, Model 6). From this analysis, we can conclude that the booster administration rate was very important in preventing deaths during the period when Omicron BA.1/BA.2 dominated.

### Comparison of COVID-19-related mortality in “faster” and “slower” countries during the dominance of different SARS-CoV-2 variants

Four epidemic waves caused by Wuhan, Alpha, Delta, and Omicron viruses BA.1, as well as BA.2, resulted in different excess mortality rates in different countries, as shown in Figure 3B. We were interested in knowing how the magnitude of excess mortality in different periods of the pandemic depended on which category a country was in, namely whether it was “faster” or “slower” in its vaccination.

To answer this question, the distributions of excess mortality values were visualized as box plots for both categories of countries for the three intervals of the pandemic (Figure 8 A). The first interval (I) included the time prior to mass vaccination. The second (II) and third (III) periods correspond to the time when vaccination was in full swing. As mentioned above, viral variants Wuhan and Alpha (pre-Delta strains) dominated in the first interval. Delta in the second interval, and Omicron BA 1/BA.2. in the third. We then assessed the ratio of averages in each data category. By comparing these averages, we obtained information on how the probability of dying during each pandemic period was related to the category of country, in which the individual lived (Figure 8 B). A low probability of dying during a pandemic meant better protection against all-cause mortality provided by countries in the corresponding category.

**Figure 8.**
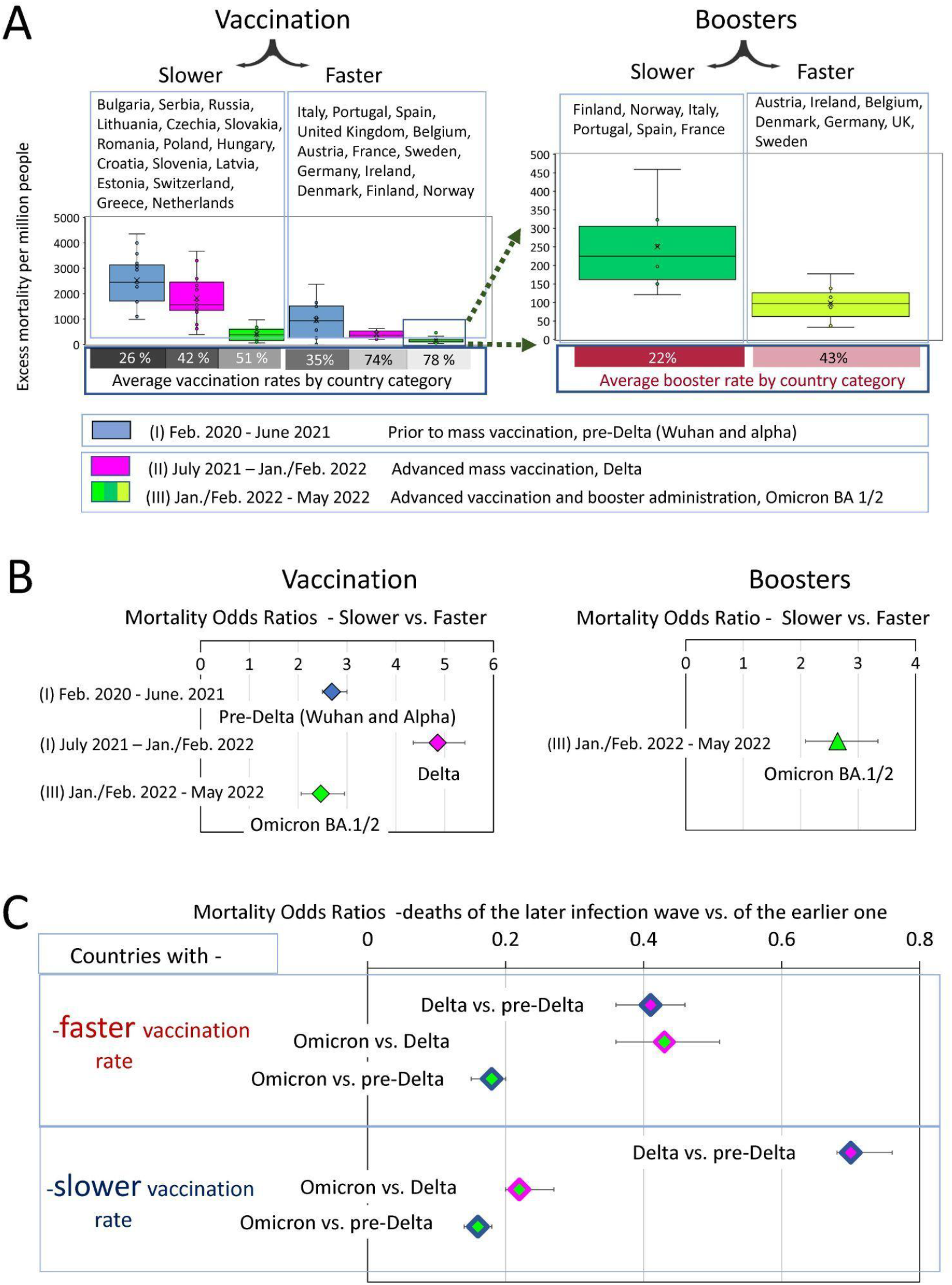
Analysis of COVID-19 associated excess deaths averaged for country categories. A. Distribution of excess mortality values in two categories of countries at three different time intervals of the pandemic, visualized as boxplots. B. Odds ratio of dying in each time interval depending on the country category. С. Odds ratio of dying in later infection waves versus the previous infection waves depending on the country category.

The “faster” countries, compared to the “slower” ones, were much better at protecting their residents throughout the pandemic. However, the difference in protection effectiveness depended on the time interval. For example, the odds of dying in the first pre-Delta time interval were nearly three times higher for residents of countries that failed to provide prompt vaccination in the future compared with those that did (OR 2.9 (95% CI 2.5-3)). The contrast in the odds of dying between these same categories of countries became much more pronounced, reaching almost fivefold in the second time interval, when vaccination was advanced, and the Delta variant dominated (OR 4.9 (95% CI 4.4-5.4)). However, the contrast decreased twofold in the third time interval, when people died from the Omicron variant to an OR of 2.5 (95% CI 2.1-2.9). For the same wave of Omicron infections, there was more than a twofold difference in mortality for the “boosters slower” versus “boosters faster” subcategory with an OR of 2.6 (95% CI2.1-3.3).

### The degree of change in mortality during successive waves of COVID-19 infections depends on the magnitude of mortality in the previous wave and the level of vaccination

How much does the excess mortality in each successive infectious wave change from the previous wave, depending on the level of vaccination in the country? How much has the excess mortality rate changed in each successive infectious wave relative to the previous wave, depending on the vaccination rate in the country? To answer these questions, we visualized the distributions of excess mortality values as box plots (Figure 7A).

To evaluate the dynamics of mortality-change in transitions between the different COVID-19 pandemic waves, we measured odds ratios between mortality in the late and early infectious waves for “faster” and “slower” vaccinated countries separately. Thus, we estimated mortality odds ratios between Delta and pre-Delta, or Omicron and Delta waves.

The comparative analysis of the odds ratio in Figure 7C demonstrated that the first transition was much more pronounced for the “faster” countries compared to the “slower”. Mortality during the Delta wave was less than half of pre-Delta mortality in the “faster” category (OR 0.4 (95% CI 0.36-0.46)) and more than half in the “slower” category (OR 0.7 (95% CI 0.68-0.76). In other words, during the transition to the Delta wave, mortality rates declined strongly in the “faster” countries and weakly in the “slower” ones. In contrast, the transition between Delta and Omicron waves was more pronounced for the “slower” countries. In this category, Omicron-associated mortality was only one fifth of that of the Delta wave (OR 0.22 (95% CI 0.2-0.25)). In contrast, in the “faster” category, Omicron accounted for just under half of the Delta deaths (0.43 (95% CI 0.36-0.53)). Nevertheless, the odds ratios of pre-Delta versus Omicron deaths represent similar values in both categories of countries (Figure 7C).

Thus, we can summarize that the degree of mortality reduction during the transition from infection waves caused by pre-Delta virus variants to the Omicron variant was independent of the rate of vaccination in the countries. However, the trajectory of this decrease depended on this rate. We have seen a sharp decline between pre-Delta and Delta mortality for the “faster” countries. At the same time, we observed a strong mortality reduction between Delta and Omicron waves for the “slower” countries. The difference in trajectories led to the major difference in mortality between the two categories of countries during the Delta infection wave. The ability of rapid vaccination to save lives was best exemplified by the Delta wave, and the ability of rapid booster administration to save lives was best exemplified by the first Omicron wave.

## Discussion

### Estimates of excess mortality versus COVID-19 confirmed deaths

The underreporting of COVID-19 deaths in some countries is a well-known phenomenon, which was thoroughly discussed in previously published studies (14). The global worldwide estimate is 18 million excess deaths between early 2020 and the end of 2021, while reported COVID-19 deaths over the same period are about 6 million, three times less (37).There are several reasons for the discrepancy between reported and excess COVID-19 deaths. For example, medical reporting systems may not list COVID-19 as a cause of death if a person has not been tested for SARS-CoV-2, and thus deaths caused by the virus may be missed in official counts in countries with low testing rates. Early in the pandemic, before widespread testing, many COVID-19 deaths among the elderly were not related to the disease, causing a significant underreporting in some countries (16).

Thus, our study found that in some countries there is a disparity between excess mortality and deaths directly related to COVID-19 which is not unexpected. The existence of this discrepancy, and the fact that it occurs primarily in countries with relatively low GDP per capita, is consistent with what has already been found and published (14, 17). In summary, all these findings underscore the fact that excess mortality is a more reliable indicator of pandemic deaths than COVID-19 direct mortality, which has been diagnosed as a direct result of COVID-19.

### Low GDP, low vaccination rates and high pandemic-associated mortality

Our study showed that countries with low GDP per capita have higher mortality rates. A negative correlation between excess mortality and GDP per capita has been observed before. It has been seen for Spanish flu (26) and for COVID-19 pandemic (27, 28). Even at the single-country level, the excess mortality associated with COVID-19 is inversely correlated with the average family income that existed in the area of residence (25). Thus, our observations are consistent with those found earlier in published studies. Not surprisingly, richer countries have more resources to deal with the pandemic-induced problems, so they do a better job of reducing excess mortality overall.

Also not surprisingly, our analysis demonstrated a positive correlation between GDPs per capita and vaccination rates. The data showed that countries with relatively low incomes were slower to vaccinate their citizens and ended up with lower vaccinated populations. Similar observations have been described in detail in the research publications (19, 29). It has also previously been observed that low vaccination rates in countries coincide with underreporting of COVID-19 mortality (18). In this context, the results of our analysis of European countries, which show a discrepancy between COVID-19 and excess mortality as well as low rates of vaccination in countries with low GDP, are consistent with previous findings.

### Age, life expectancy and excess mortality

COVID-19 is more dangerous for elderly people, and deaths occur primarily in the older population (38). In our analysis, however, we found only a weak relationship between the average age of people in a country and excess mortality (Figure 6A). The correlation did not reach statistical significance and was detected only in those countries where vaccination was faster (Supplementary Figure 3, lower left panel). Also we didn’t find any correlation between percentage 65+ people in a country and excess mortality (Figure 6B and supplementary Figure 3, middle panels.) This may indicate that the level of medical care and vaccination rate play a greater role in saving lives than the average age of the population or percentage of elderly population in a country. This conclusion is also supported by the strong negative correlation between life expectancy and excess mortality (Figure 6C). The higher the life expectancy, the fewer lives the country lost during the pandemic. This is true for both the Delta wave and the Omicron wave.

### Finding a causal relationship between COVID-19 pandemic mortality and rates of vaccination and booster

Analysis of pandemic mortality across countries allows us to examine the effectiveness of vaccines and boosters during different periods of the pandemic when different variants of the coronavirus were prevalent. In addition, such analyses allow an assessment of how well vaccination worked against the background of immunity triggered by natural infections. The significant and strong negative correlations we observed between national vaccination rates and excess mortality seemed to answer the question of vaccination effectiveness in reducing COVID-19 mortality in an obvious way. However, a closer analysis of the data showed that other factors, namely public health effectiveness, quality of healthcare, and the efficacy of pandemic mitigation strategies, must also be considered to assess the impact of vaccination on saving lives. Isolation of the impact of these factors is not a straightforward task. They are all linked and act synergistically.

In our work, we analyzed the data to distinguish the contribution of these listed factors from the vaccination rate factor. In doing so, we assumed that annual actual GDP per capita largely determines the amount of funding available to national governments to implement all life-saving strategies, including those not related to vaccination rates.

Our analysis shows that “faster” countries that achieved higher vaccination rates had lower pre-vaccination excess mortality compared to countries with low vaccination rates. However, the difference between “faster” and “slower” countries became much more pronounced when mass vaccination was in full swing. From this we conclude that although countries differed in the effectiveness of COVID-19 mortality control measures before vaccination, vaccination made these differences much more pronounced. Thus, vaccines greatly improved the effectiveness of pandemic control measures.

In this study, we found the existence of a certain threshold level of vaccination, namely 60-70% of the country’s population. Countries that reach this threshold quickly differ little in their mortality rates in comparison to the slower vaccinating countries, where the difference was significant. However, during Omicron dominance, despite the threshold reached, the countries that reached it, still differed in terms of excess mortality, and the magnitude of this excess mortality correlated inversely with the level of booster vaccinations. The immuno-compromising characteristics of Omicron likely contributed to diminished protective effect of the vaccination.

Our work pointed to the great importance of rapid administration of boosters before January 2022. Mortality in countries with rapid booster administration was significantly lower than in countries with the same per capita GDP, the same vaccination rate, but lower booster rates. The results of our data analysis are consistent with the observation that additional booster doses of both mRNA- and adenovirus-vector-based vaccines significantly increase the protective efficacy of vaccine against severe disease (13).

### The transition from one infectious wave to another and the associated change in mortality in “faster” and “slower” countries

In our work, we have shown that the overall rate of COVID-19 related deaths varies across countries and depends on many factors, including the level of vaccination. However, even before vaccination, countries in the category where the population would subsequently be vaccinated more quickly had lower excess mortality rates. Apparently, this is due to the fact that these countries have on average, higher GDP per capita and, accordingly, more capacity to mitigate the epidemic consequences. At the same time, during the period of mass vaccination, and especially during the period of dominance of the Delta variant, the ability to reduce mortality increased sharply in the category of countries that were rapidly vaccinating their population. The contrast in terms of excess mortality between rapidly or slowly vaccinating countries became particularly strong. Accordingly, the inverse correlation between the number of vaccinated people in the country and excess mortality became particularly pronounced during the Delta infection wave. This correlation weakened in the next infectious wave, namely, the Omicron-dominated wave. In fact, the difference between fast and slow vaccinating countries became the same as it was before mass vaccination began. There are several explanations for this phenomenon. First, the time has passed since the vaccination and the immune vaccine defense has weakened. Second, Omicron has a more pronounced ability to resist immune defenses, and finally, immune defenses increased after natural infections in countries with low vaccination rates.

It is worth noting that booster vaccinations played a hugely positive role in reducing mortality during the Omicron BA.1/BA.2 dominated period. In countries with comparable levels of GDP per capita and similar rates of primary series vaccination, excess mortality was largely determined by the level of booster vaccination administration during the Omicron-dominated period. This observation tells us that even though Omicron antigenically is very different from the ancestral SARS-CoV-2 strain, for which the vaccines were produced, booster vaccination effectively prevented excess mortality in the first waves of Omicron, caused by BA.1 and BA.2 virus variants.

Our work revealed an interesting pattern, namely that the degree of reduction in excess mortality when comparing the pre-Delta to Omicron waves was independent of the rate of vaccination. In Figure 7C, we showed that Omicron mortality is about one-sixth that of pre-Delta mortality in both the “faster” and “slower” categories. However, mortality decreases equally only in the transition from the pre-Delta wave to the Omicron one. As for the intermediate transitions, they are very different in the two categories of countries. The most pronounced reduction in excess mortality occurs in the transition to Delta in the category of countries with rapid vaccination and in the transition to the Omicron wave in the category of countries with slower vaccination rates.

A possible explanation is that rapidly vaccinated countries developed immunity faster, mostly in the pre-Delta pandemic phase, and slow-vaccinated countries developed hybrid (vaccine plus disease) immunity in later phases, likely during the Delta wave of infection. Thus, immunity, which saves from death, developed more rapidly in some countries, mainly due to vaccination, and more slowly in others, because of the hybrid influence of vaccine administration and natural infections. Despite all this, however, the minimum number of deaths in the first wave of Omicron was in countries with the highest rates of booster vaccination. Our results demonstrated that booster protection can still have a significant impact and reduce excess mortality despite high levels of immunity in the populations of countries in both categories.

### Hybrid immunity

Excess mortality during the Omicron BA.1/BA.2 waves is approximately one-sixth that of pre-Delta mortality in both “fast” and “slow” countries (Figure 7C). We believe that this finding indicates that by the time the Omicron variant appeared, the general immunity of the population had already developed and could be comparable between countries from both categories. The protective shield of established immunity significantly reduced the number of deaths during the Omicron wave compared to previous periods. It has been shown that prior SARS-CoV-2 infection and booster vaccinations provide strong protection from omicron caused ICU admission or deaths (39, 40, 41). Therefore, we can assume that the immunity protective shield was formed as a result of vaccination or COVID-19 disease and, importantly, sometimes as a combination of both, in the same person, namely as a results of hybrid immunity. The latter is particularly important and has recently been shown to be able to protect more effectively than natural immunity acquired by people from vaccine alone or disease alone (42, 43, 44). It is likely that the percentage of people with hybrid immunity increased significantly in subsequent waves of Omicron variants and thereby strengthened population immunity. The existence and maintenance of population immunity can probably explain, at least in part, the absence of large infectious waves in the second half of 2022 in most countries.

Many studies have compared immunity from vaccines, natural and breakthrough infections and have shown that hybrid immunity protects better against SARS-CoV-2 variants compared to immunity from vaccine alone or COVID-19 alone (42, 43, 44). When vaccinating previously infected individuals, just one dose of vaccine enhances both B- and T-cell response to various variants of the virus (45). In individuals previously infected with COVID-19, vaccination induces the production of cross-variant neutralizing antibodies (46).

In some countries, especially in China, the formation of hybrid immunity, in particular its component derived from natural immunity against the disease, has been severely delayed. This is primarily due to a three-year policy of rigorous testing, contact tracing combined with strict quarantines and even lockdowns, commonly referred to as the “zero covid” policy. This policy was introduced in China in January 2020 and continued until December 2022 (10, 47).

Vaccine immunity is gradually lost, and the rate of loss of protective effectiveness depends on the type of vaccine. China used types of vaccines based on an inactivated virus. A comparison of these vaccines and mRNA-based vaccines used in many European countries and Hong Kong during the Omicron BA.2 outbreak in March 2022 showed that vaccines based on an inactivated virus protected people for a shorter period of time than mRNA based vaccines (10).

In addition, the percentage of the booster-vaccinated population among the elderly, who are particularly at risk for fatal or severe COVID-19, was relatively small in China. Some scientists familiar with the epidemiological situation in China have advised urgently increasing this percentage (47). Thus, the abrupt cancellation of China’s zero-covid policy in late 2022 led to a dramatic outbreak of disease, leading to a significant increase in hospitalizations in December and January. Already in the first week of January, the number of hospitalizations, according to the WHO, doubled and mortality rates also rose sharply (48).

Natural immunity to COVID-19 in Hong Kong in January 2023 was probably better than in mainland China because the island survived a severe outbreak of Omicron BA.2 in March 2022. Some experts estimated that about 40% of Hong Kong’s population was infected during the outbreak (OurworldInData, (32)). It is worth noting that the vaccination situation in Hong Kong is better than in mainland China because almost half of the population received the mRNA vaccine, which has a longer protection period than the inactivated virus-based vaccines used in China (10). Considering these data, it can be assumed that the situation in mainland China can only be worse than in Hong Kong. The daily mortality rate on the mainland seems to be reaching a higher level than on the island, where there were nine deaths per million people in January 2023. However, using Hong Kong data, even conservative estimates for China’s population of 1.4 billion predict a daily death rate of 12600.

Hybrid immunity has probably developed faster in countries that do not have such radical policies as China. It has been shown that regardless of the vaccine used, hybrid immunity induces a stronger humoral response than vaccination (44). Hybrid immunity may also provide greater protection than immunity induced by vaccination alone against the Omicron variant (49). We assume that some infectious background of continuing circulating SARS-CoV-2 variants along with booster vaccinations will maintain hybrid immunity in most countries going forward. As a result, we can expect that in 2023 most of the world, and especially the European countries considered in this study, will avoid major infectious outbreaks such as the one that occurred in China in January of this year.

## Conclusions

A. Slow vaccination and slow booster administration have been associated with high excess mortality from COVID-19 in European countries. In contrast, high vaccination rates provided robust protection against virus-associated mortality. Vaccine protection peaked in the Delta wave but became weaker in the Omicron wave.
B. However, additional booster vaccination was very effective in preventing excess mortality caused by the Omicron BA.1/BA.2 infectious wave.
C. The main trend found in this study was that the European countries that vaccinated their populations faster were mostly the same countries that had higher GDP per capita. They also provided better protection against COVID-19-related deaths even before vaccination campaigns began.
D. Although a small number of countries protected their populations from COVID-19 deaths poorly before vaccination campaigns began, they did much better afterwards by ensuring fast vaccination of their citizens.
E. The excess mortality during the COVID-19 pandemic correlates not only with a county’s vaccination rate, but also with its per capita GDP. The latter parameter likely reflects and is related to the quality of healthcare in the country, the availability of mass COVID-19 testing, and funding for other pandemic mitigation strategies.

## Data Availability

All data used in the study are from public resources and open-access databases.
https://ourworldindata.org/coronavirus
https://data.worldbank.org/

https://data.worldbank.org/

## Funding and acknowledgements

This work was supported by the Intramural Research Program of the National Library of Medicine, National Institutes of Health (SAS).

## Institutional Review Board Statement

The study does not fall within the regulatory definition of research involving human subjects. Current research involves only secondary data analysis of already publicly available datasets, which contain no information that can identify subjects, directly or through identifiers linked to the subjects.

## Contributions

OVM - Data curation, Formal analysis, Investigation, Figures design, Writing

SAS - Conceptualization, Methodology, Writing

## Data Availability

All data used in the study are from public resources and open-access databases.

## Figures and legends

**Supplementary Figure 1.**
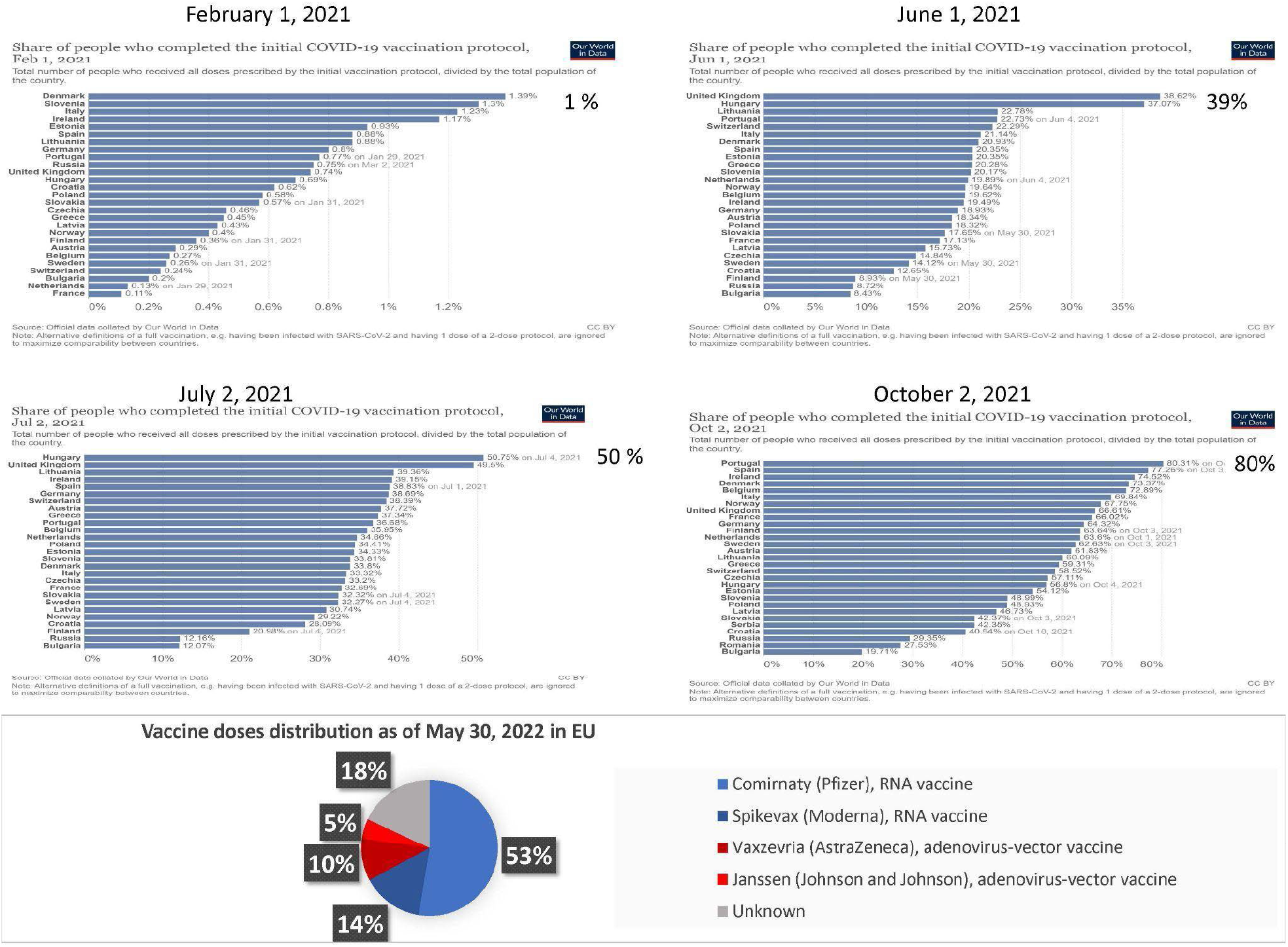
Vaccination rates and vaccine types in Europe.

**Supplementary Figure 2.**
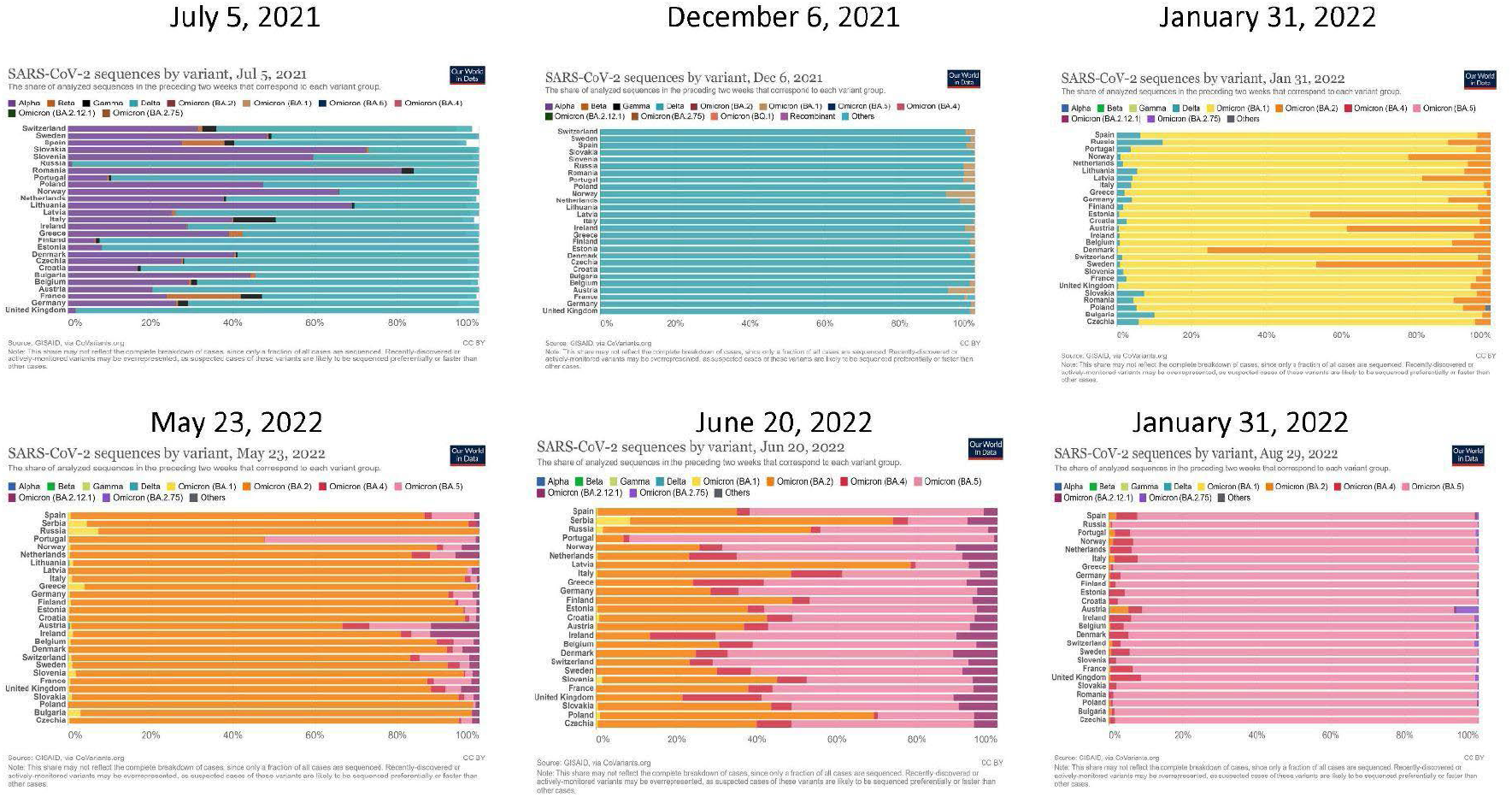
Changing dominant variants of SARS-CoV-2 in Europe.

**Supplementary Figure 3.**
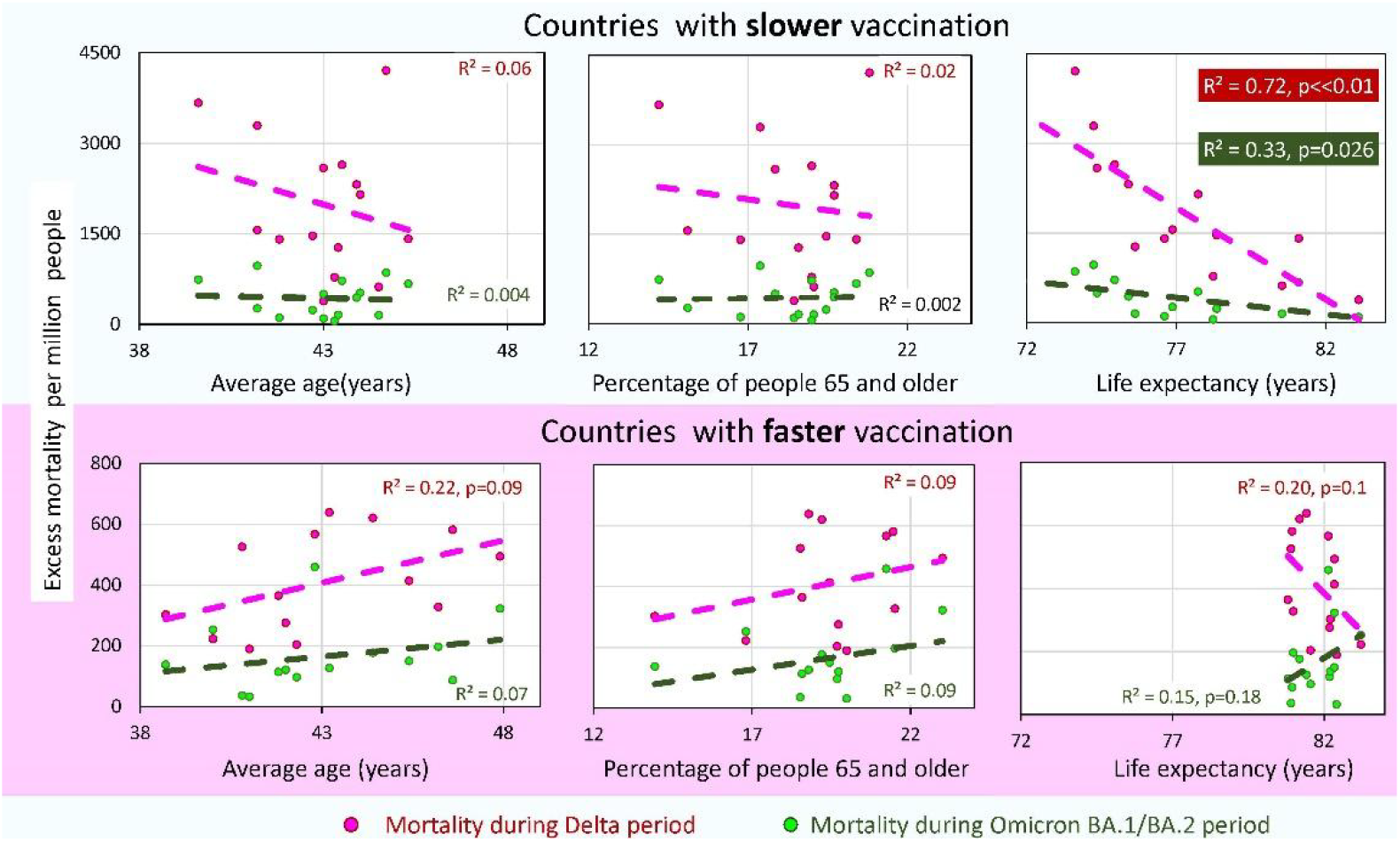
Analysis of excess mortality, the average age of the population in the country, the percentage of older people, and life expectancy in the country for two categories of countries.

